# Percentage of reported Covid-19 cases in Colombia: Estimating the true scale of the pandemic

**DOI:** 10.1101/2020.12.30.20249052

**Authors:** Nicolás Parra-A, Vladimir Vargas-Calderón, Juan Sebastián Flórez, Leonel Ardila, Carlos Viviescas

## Abstract

At the outbreak of a virus, data on cases is sparse and commonly severe cases, with a higher probability of a fatal resolution, are detected at a larger rate than mild cases. In addition, in an under-sampling situation, the number of total cases is under-estimated leading to a biased case fatality rate estimation, most likely inflating the virus mortality. In this communication, we present a method to estimate the sub-report in a country that accounts for both the delay time between symptoms onset to death and the country’s demographics. The method is based on the comparison of the corrected case fatality rate (CFR) of the target country with the one of a benchmark country. Using reported data from Instituto Nacional de Salud up to December 28, we utilize our method to provide a comprehensive estimate of the Covid-19 sub-report in Colombia, its regions and some of its cities during 2020.

## I. INTRODUCTION

Since March 6, when Colombia confirmed its first case of the coronavirus disease (Covid-19), the country healthcare system, with a limited testing capability, has struggled to monitor and report current cases. Keeping track of active cases in a pandemic is of paramount importance for epidemiological tracing in the early stages of the contagion spread. In more advanced stages of a pandemic, when epidemiological routes cannot be constructed, effectively testing the population becomes a challenging task that must thrive to keep the cases reported as high as possible. The accuracy in measuring case incidence and prevalence, as well as mortality rates is decisive for high-quality epidemiological modelling that enables governments to propose and implement public policies to mitigate the impact of the pandemic [1–3].

Throughout the current Covid-19 pandemic, governments have compiled extensive databases with information on infected people. In particular, data on positive cases and deaths are relevant for tracking the infection evolution. Commonly, however, tests are performed only on a subset of the total country’s population. How this subset is selected varies according to the country’s resources and the chosen strategy, but no matter how effective this strategy turns out to be, there are always some infected individuals that do not ever get tested for the disease and, therefore, are not registered as a positive Covid-19 case in the database. This phenomenon is known as under-reporting, and its magnitude varies especially in relation with the effectiveness of epidemiological tracing strategies, as well as tests availability [4].

In this work we propose a method to estimate the sub-report in a country from disease case databases. The value of the percentage of cases reported found in this way can then be used to obtain better quality figures for the incidence of cases and mortality rates. In particular, we analyze the data reported for Colombia, one of the most affected countries by the pandemic, its departments and some of its regions during 2020. Unlike seroprevalence studies, the proposed method needs not perform further tests than the ones already conducted, and whose results are contained in the cases databases.

Central to our method is the comparison of the case fatality ratio (CFR) of a target country, which in our case is Colombia, to the CFR of a benchmark country, which in this work is the Republic of Korea. A criterion for its selection as a benchmark, is that the track and testing strategy followed by the country has been effective, leading to a low under-reporting of Covid-19 cases. In order to account for differences between the population age distribution of the target and the benchmark country our method also incorporates the demographics of the countries. In this sense, the method answers the question of how many positive cases would be detected in the target country if it were using the testing strategy of the benchmark country?

The outline of the paper is as follows. The details of the method as well as a description of the data used are presented in Section II. Our estimates of the sub-report of Covid-19 for Colombia and its regions are reported in Section III, and are further discussed in section IV, where we also compare our work with other studies. The limitations of our approach are listed in section V. Finally, we conclude in section VI.

## II. METHODS AND MATERIALS

### A. Data

We use two datasets for our analysis. The first one is the data published by the Instituto Nacional de Salud (INS) [5], which is updated every day and reports all known Covid-19 cases in Colombia. For each case it provides its date of notification, location city, department or district, current status (recovered, recovery from home, being treated in an intensive care unit and passed away), age, sex, country of provenance, symptom onset date, date of death, date of diagnosis, date of recovery and the web report date. To construct the case and death incidence time-series (see fig. 1) we used data up to December 28, 2020. In addition, we also keep information about age and location of each case.

**FIG. 1.**
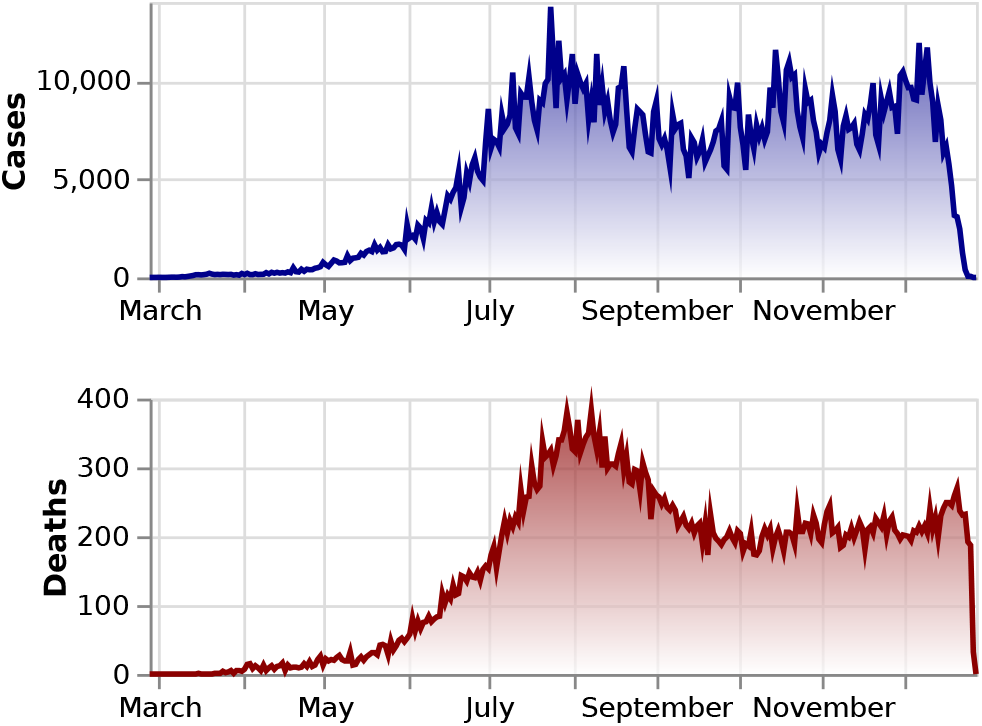
Covid-19 cases and deaths incidence in Colombia. Up to December 28, 2020 there have been 1’594,497 confirmed cases and 47,175 confirmed deaths.

The other dataset is extracted from the demographic database maintained by the Departamento Administrativo Nacional de Estadística (DANE) [6]. We retrieve data containing the most recent projection for the year 2020 of the number of individuals per age group for all regions and cities of Colombia (see fig. 2). We make use of this information to assess the vulnerability of each region to Covid-19.

**FIG. 2.**
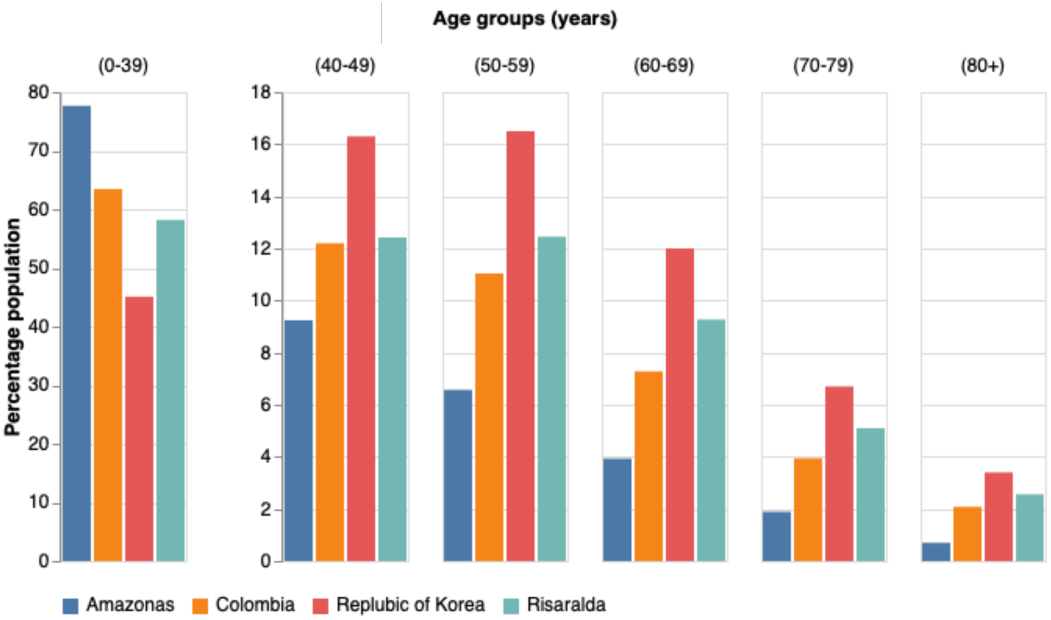
Population demographic comparison by age-group between Amazonas (one of the youngest populations in Colombia), Risaralda (one of the oldest populations in Colombia), Colombia and the Republic of Korea.

### B. Delay adjusted case fatality rate estimate

Dividing deaths-to-date by cases-to-date to compute the CFR (from now on, nCFR indicates this naïve determination of the CFR) tends to yield a biased result, usually underestimating its true value [7, 8]. This happens because the outcome of active cases (i.e. recovery or death) is still not known. An improved calculation of the CFR can be done by accounting for the delay from onset-to-death to estimate the number of cases expected to have a known outcome.

#### 1. Evaluating the delay time distribution between onset-to-death

We used the reported dates to calculate the time interval from illness onset to death of the confirmed 29,480 cases that resolved in death from March 16 to September 19 2020 in Colombia. The totality of cases in this period were already resolved (i.e. recovered or dead). We fit the conditional probability density *f* (*t*) of the time between onset-to-death given death to Weibull, gamma, and lognormal distributions [9, 10]. In all cases we use the maximum likelihood estimation (MLE) to determine the parameters and obtain confidence intervals using PyMC3 [11]. In addition, we include a gaussian kernel density estimation (KDE). We select the best fit model by using the Akaike information criterion (AIC). Table I shows estimates for the three models plus the KDE. In fig. 3 we show our gaussian KDE approximation to the onset-to-death distribution, with a mean time of 22.4 days and a standard deviation of 12.7 days. Our results for Colombia differ with distributions that were reported earlier in the pandemic [9, 10].

**TABLE I.**
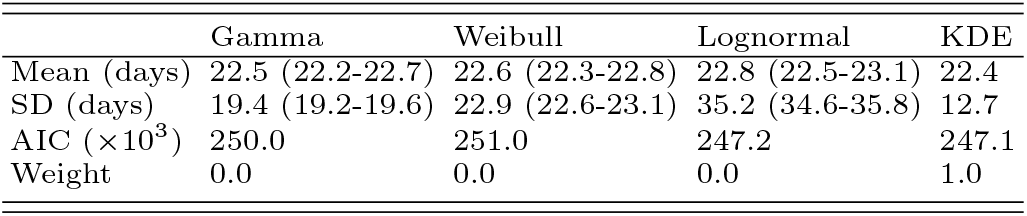
Illness onset to death time-delay distribution for Covid-19 outbreak in Colombia. 95% confidence intervals are displayed inside parenthesis.

|  | Gamma | Weibull | Lognormal | KDE |
| --- | --- | --- | --- | --- |
| Mean (days) | 22.5 (22.2-22.7) | 22.6 (22.3-22.8) | 22.8 (22.5-23.1) | 22.4 |
| SD (days) | 19.4 (19.2-19.6) | 22.9 (22.6-23.1) | 35.2 (34.6-35.8) | 12.7 |
| AIC ($\times 10^3$) | 250.0 | 251.0 | 247.2 | 247.1 |
| Weight | 0.0 | 0.0 | 0.0 | 1.0 |

**FIG. 3.**
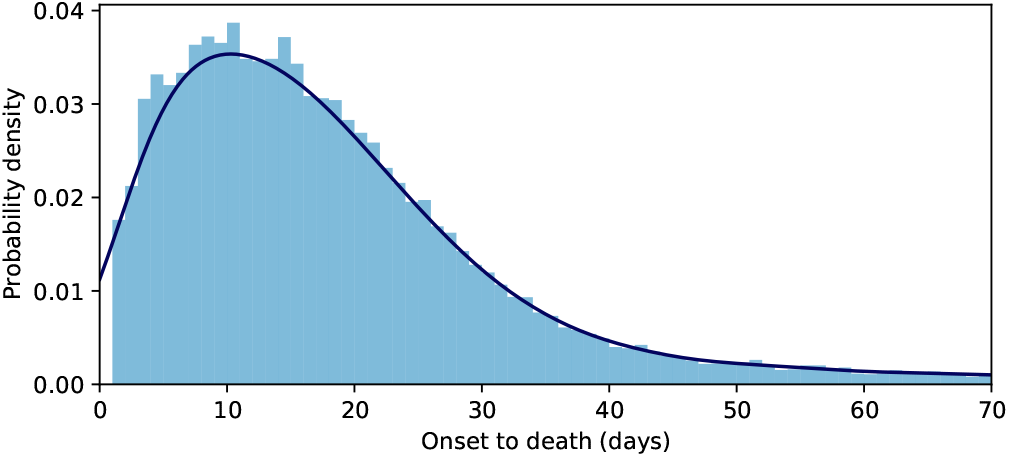
Probability density distribution of the time from illness onset to death *f* (*t*). The continuous line corresponds to a KDE fit with mean delay of 22.4 days and standard deviation of 12.7 days.

#### 2. Adjusted case fatality ratio

The CFR is adjusted following the method proposed by Nishiura *et al*. [7]. We estimate the proportion of cases resolved using the case and death incidence to modify the CFR in order to account for delay outcome (cCFR). The underestimation factor [7, 8, 12–14],

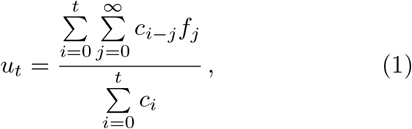

scales the cumulative number of cases in the denominator of the cCFR, and makes up for the adjustment. Here *c*_*t*_ is the daily case incidence (see top panel of fig. 1) at time *t* and *f*_*t*_ = *f* (*t*) is the conditional probability density of the delay-time from onset-to-death (see fig. 3).

The severity of the Covid-19 is highly correlated to the age of the infected individual [10], hence, for each region or city, we evaluate age-stratified estimates of the adjusted case fatality rate, 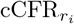, where *r* stands for the region and *i* labels the age group. The age aggregated cCFR for a region is adjusted for the population demographics,

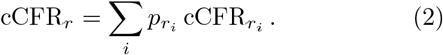

Here,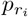 is the fraction of the population with age *i* for the region *r*.

### C. Percentage of cases reported

The adjusted cCFR does not account for under-reporting. In order to obtain an estimate of the potential level of under-reporting in Colombia and its regions our model follows the method proposed by Russell *et al*. [12], further adjusting for the demographics of the country.

We assume a baseline CFR (blCFR), taken from a benchmark country, and compare it with the estimated cCFR for Colombia and some of its regions and cities. We do an age-stratified analysis. If the 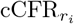 is higher than the age-group baseline, blCFR_*i*_, it indicates that only a portion of the real number of cases in this age group have been reported so far. The fraction of reported cases in region *r* and age group *i* is given by

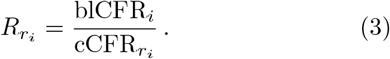

We also evaluate the fraction of cases reported in a region aggregated over age *R*_*r*_. For this, we introduce the region baseline CFR,

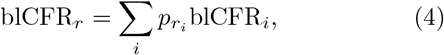

which accounts for the region population demographics through 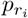, and obtain

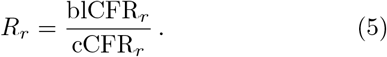

The most recent literature offers several estimates of CFR for Covid-19. Among those adjusting or controlling for under-reporting, we mention those by Verity *et al*. [10], Russell *et al*. [13], Shim *et al*. [14], Guan *et al*. [15]. In our analysis, we use as benchmark country the Republic of Korea. As baseline for this work we take the age stratified CFR data of July 14, 2020 [16], listed in Table II, since most of the cases from the first peak had already been resolved by this date.

**TABLE II.** Covid-19 case fatality rates stratified by age groups as of July 14, 2020 in the Republic of Korea.

| Age Groups (years) | Case Fatality Ratio (95% CI) |
| --- | --- |
| 0-39 | 0.04 % (0.00-0.09) |
| 40-49 | 0.17 % (0.00-0.36) |
| 50-59 | 0.62 % (0.31-0.93) |
| 60-69 | 2.32 % (1.62-3.02) |
| 70-79 | 9.31 % (7.42-11.21) |
| ≥80 | 24.96 % (21.43-28.49) |

## III. RESULTS

In this study we consider all regions and cities in Colombia that have reported at least 40 fatal Covid-19 cases as of December 28, 2020. Table III shows the percentage of cases reported, the cCFR, blCFR and the total cases and deaths for all regions and some cities of Colombia. For the country the blCFR obtained is 1.2% (95% CI: 0.9-1.5) while its cCFR value is 2.7% (95% CI: 2.7-2.8) leading to a percentage of cases reported of 43% (95% CI: 43-43). It is worth remarking that Colombia’s blCFR is lower than the Republic of Korea’s, 1.9% (95% CI: 1.5-2.3) [17], due to its younger population. At the level of regions and cities these values fluctuate appreciably.

**TABLE III.** Percentage of Covid-19 cases reported in Colombia and its regions until December 28, 2020 with 95% confidence intervals. The corrected and baseline CFRs are also shown, along with the total number of positive cases and deaths to date. Regions are numbered to later compare with fig. 4 (N.S. identifies regions that are not shown in fig. 4 because they are districts and not a departments).

|  | Percentage<br>of cases reported | cCFR | blCFR | Total cases | Total deaths |
| --- | --- | --- | --- | --- | --- |
| Colombia | 43 % (43-43) | 2.7 % (2.7-2.8) | 1.2 % (0.9-1.5) | 1594497 | 47175 |
| AMAZONAS (1) | 24 % (20-28) | 2.2 % (1.4-3.3) | 0.5 % (0.4-0.7) | 3231 | 134 |
| ANTIOQUIA (2) | 56 % (55-58) | 2.2 % (2.1-2.3) | 1.2 % (0.9-1.5) | 255845 | 5220 |
| ARAUCA (3) | 31 % (26-36) | 2.4 % (1.6-3.5) | 0.7 % (0.5-1.0) | 4521 | 150 |
| ATLANTICO (4) | 29 % (27-30) | 3.8 % (3.4-4.2) | 1.1 % (0.8-1.4) | 37024 | 1690 |
| BARRANQUILLA (N.S.) | 36 % (35-38) | 3.5 % (3.1-3.8) | 1.3 % (1.0-1.6) | 54725 | 2082 |
| BOGOTA (5) | 48 % (47-49) | 2.3 % (2.3-2.4) | 1.1 % (0.9-1.4) | 452940 | 10865 |
| BOLIVAR (6) | 37 % (33-41) | 2.8 % (2.2-3.7) | 1.0 % (0.8-1.3) | 8124 | 333 |
| BOYACA (7) | 55 % (51-59) | 2.7 % (2.2-3.1) | 1.5 % (1.1-1.8) | 27620 | 681 |
| CALDAS (8) | 66 % (62-71) | 2.3 % (2.0-2.7) | 1.5 % (1.2-1.9) | 31604 | 765 |
| CAQUETA (9) | 30 % (28-32) | 2.8 % (2.3-3.3) | 0.8 % (0.6-1.0) | 14845 | 599 |
| CARTAGENA (N.S.) | 57 % (53-61) | 1.8 % (1.5-2.1) | 1.0 % (0.8-1.3) | 42318 | 819 |
| CASANARE (10) | 33 % (29-38) | 2.2 % (1.5-3.0) | 0.7 % (0.5-0.9) | 8578 | 197 |
| CAUCA (11) | 47 % (44-51) | 2.4 % (2.0-2.9) | 1.1 % (0.9-1.4) | 18174 | 578 |
| CESAR (12) | 34 % (32-36) | 2.4 % (2.1-2.7) | 0.8 % (0.6-1.0) | 33161 | 1117 |
| CHOCO (13) | 28 % (25-33) | 2.7 % (1.9-3.7) | 0.8 % (0.6-1.0) | 4800 | 199 |
| CORDOBA (14) | 27 % (26-28) | 4.0 % (3.6-4.4) | 1.1 % (0.8-1.4) | 29427 | 1892 |
| CUNDINAMARCA (15) | 42 % (40-44) | 2.8 % (2.5-3.1) | 1.2 % (0.9-1.5) | 64803 | 1877 |
| GUAINIA (16) | 35 % (24-53) | 1.2 % (0.3-3.1) | 0.4 % (0.3-0.6) | 1229 | 23 |
| GUAJIRA (17) | 29 % (26-31) | 2.2 % (1.8-2.7) | 0.6 % (0.5-0.8) | 13938 | 563 |
| GUAVIARE (18) | 36 % (27-49) | 1.8 % (0.9-3.5) | 0.7 % (0.5-0.9) | 1939 | 40 |
| HUILA (19) | 37 % (35-39) | 3.0 % (2.6-3.4) | 1.1 % (0.8-1.4) | 34205 | 1190 |
| MAGDALENA (20) | 21 % (19-23) | 4.4 % (3.6-5.4) | 0.9 % (0.7-1.2) | 7525 | 575 |
| META (21) | 39 % (37-42) | 2.3 % (2.0-2.7) | 0.9 % (0.7-1.2) | 33144 | 830 |
| NARIÑO (22) | 39 % (37-41) | 3.2 % (2.8-3.7) | 1.3 % (1.0-1.6) | 30289 | 1064 |
| NORTE SANTANDER (23) | 24 % (23-25) | 4.4 % (3.9-4.8) | 1.0 % (0.8-1.3) | 38993 | 2048 |
| PUTUMAYO (24) | 24 % (21-27) | 3.3 % (2.5-4.4) | 0.8 % (0.6-1.0) | 5791 | 258 |
| QUINDIO (25) | 54 % (50-58) | 2.9 % (2.5-3.4) | 1.6 % (1.2-2.0) | 22919 | 671 |
| RISARALDA (26) | 53 % (50-57) | 2.7 % (2.3-3.2) | 1.4 % (1.1-1.8) | 32719 | 813 |
| SAN ANDRES (N.S.) | 78 % (58-100) | 1.2 % (0.6-2.3) | 0.9 % (0.7-1.2) | 2431 | 40 |
| SANTANDER (27) | 38 % (36-39) | 3.3 % (3.0-3.6) | 1.2 % (1.0-1.6) | 65278 | 2530 |
| STA MARTA D.E. (N.S.) | 35 % (32-38) | 2.6 % (2.1-3.1) | 0.9 % (0.7-1.2) | 16760 | 583 |
| SUCRE (28) | 39 % (37-42) | 2.8 % (2.4-3.3) | 1.1 % (0.8-1.4) | 17377 | 748 |
| TOLIMA (29) | 46 % (43-48) | 3.2 % (2.9-3.6) | 1.5 % (1.1-1.8) | 42407 | 1336 |
| VALLE (30) | 41 % (40-43) | 3.5 % (3.3-3.7) | 1.4 % (1.1-1.8) | 133536 | 4636 |
| VAUPES (31) | 63 % (38-100) | 0.8 % (0.2-2.3) | 0.5 % (0.3-0.6) | 1124 | 14 |
| VICHADA (32) | 58 % (35-96) | 0.8 % (0.3-1.6) | 0.5 % (0.3-0.6) | 1136 | 15 |

Because the severity of Covid-19 varies for different age groups [10], the blCFR reflects the differences in population demographics between regions. Regions and cities in which the fraction of the population with age above 60 years is more significant display a higher blCFR compared to the value for the country. On the contrary, in those regions or cities with a younger population than the average of the country the blCFR is lower than the country’s value. We illustrate this in fig. 2, where the population age distribution of Risaralda, the department with the oldest population, Amazonas, the department with the youngest population, and the country are compared. The corresponding blCFRs mirror the demographics, Amazonas’ blCFR is 0.5% (95% CI: 0.4-0.7) while Risaralda’s is 1.4% (95% CI: 1.1-1.8). These trends are followed in specific regions of the country. Hence, Arauca (3), Caquetá (9), Casanare (10), Guainía (16), Guaviare (18), Putumayo (24), Vaupés (31) and Vichada (32) located to the east and southeast of the country, in the region of the eastern plains and the Amazon, are sparsely populated areas with a young population and a low blCFR (cf.Table III and right panel of fig. 4). In contrast, departments such as Boyacá (7), Caldas (8), Quindío (25), Risaralda (26), Tolima (29) and Valle (30), located in the densely populated Andean region, and having a considerable population of elder people, present a high blCFR (cf.Table III and right panel of fig. 4).

**FIG. 4.**
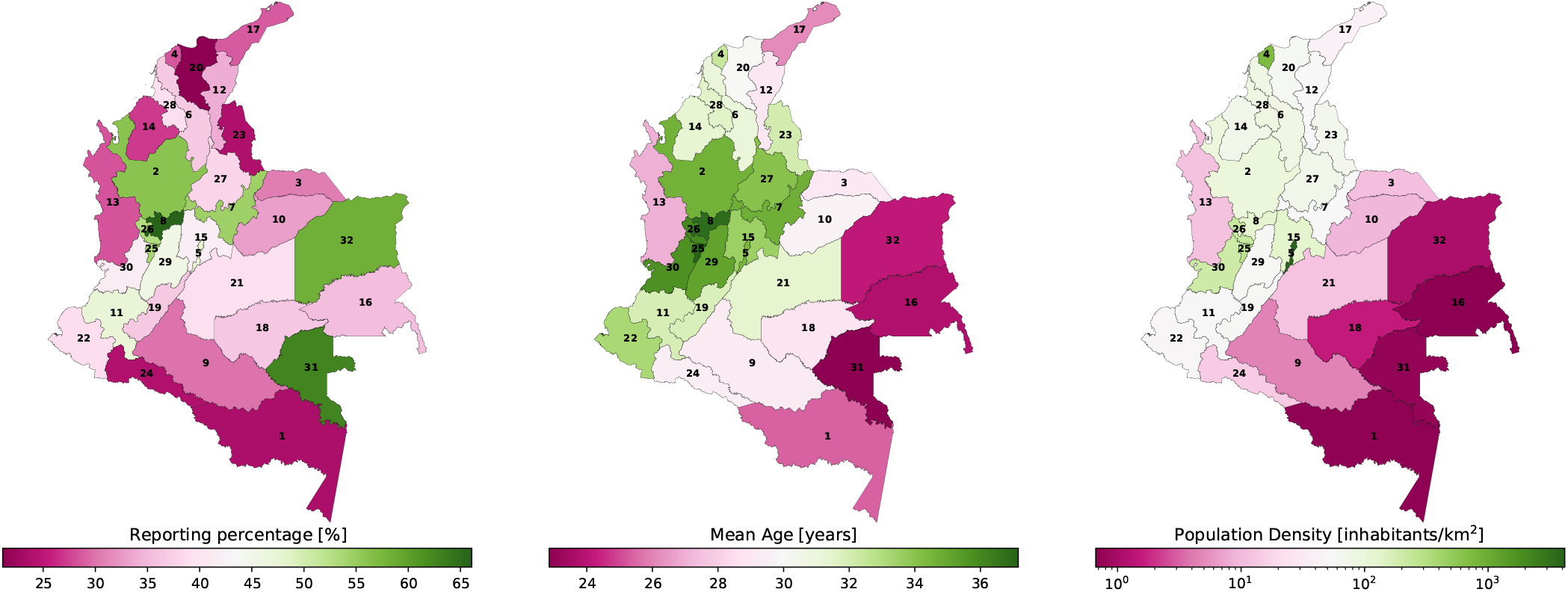
Maps of the departments of Colombia, coloured by reporting percentage (left panel), mean age (central panel), and population density (right panel).

The department with the highest percentage of cases reported are San Andrés, 78% (95% CI: 58-100), and Caldas (8), 66% (95% CI: 62-71). They are followed by Vaupés (31), 63% (95% CI: 38-100), which is the department with the smallest number of reported cases and deaths. In contrast, Magdalena (20) with a value of 21% (95% CI: 19-23), and the largest cCFR, 4.4% (95% CI: 3.6-5.4), is the department with the lowest percentage of cases reported. Bogotá (5), the capital city, where 28% of the reported cases in the country are concentrated, has a reporting percentage of 48% (95% CI: 47-49), pulling the country’s percentage of report up. Furthermore, our results show that the country can be clearly divided in regions were the reporting percentage is above or below the country’s value, revealing places where the infection has been better tracked than in others, as shown in the left panel of fig. 4. Thus, in the Andean region, where most of the country population inhabit, the majority of the departments have a percentage of cases reported above 48%. Besides Caldas (8) and the city of Bogotá (5), these also include Antioquia (2), Boyacá (7), Quindío (25), Risaralda (26), and Tolima (29). Away from the center of the country most of the regions have a low reporting percentage. Conspicuously, all departments in the Atlantic coast region have a value below 40%. To this group belong the departments of Atlántico (4), Bolívar (6), Cesar (12), Córdoba (14), Guajira (17), Magdalena (20), and Sucre (28), as well as the cities of Barranquilla and Santa Marta. An important exception is the city of Cartagena, with a reporting percentage of 57% (95% CI: 53-61); one of the highest in the country. The same behaviour is observed in the east and southeast of the country, where the departments of Amazonas (1), Arauca (3), Caquetá (9), Casanare (10), Guainía (16), Guaviare (18) and Meta (21) have a reporting percentage below 40%. In this region only the departments of Vaupés (31) and Vichada (32) set themselves apart with reporting percentages above 57%.

We present age-stratified reporting percentages in Table IV. An overall trend is noticeable: the percentage of cases reported is higher for the elder population than for the younger population (cf. central panel of fig. 4). At the country level, the percentage of reporting for the age-group > 80 is 75% (95% CI: 65-87), with all the regions and cities, except Magdalena, 50% (95% CI: 42-60), with a percentage of reporting higher than 60%. For the age group of 70-79 years, the percentage of cases reported in the country is 49% (95% CI: 40-60), with a distinct gap of more that 20 percentage points with all other age groups of younger population, and percentages of report below 15% for the groups of age below 50 years, which account for more than 70% of the country total population (see fig. 2).

**TABLE IV.** Age-stratified percentage of Covid-19 cases reported in Colombia until December 28, 2020. For the country and its regions the age-stratified percentage of reported cases are shown with a 95% confidence interval.

|  | Percentage of cases reported by age group (years) |  |  |  |  |  |
| --- | --- | --- | --- | --- | --- | --- |
| | 0-39 | 40-49 | 50-59 | 60-69 | 70-79 | $\geq 80$ |
| Colombia | 13 % (4-49) | 15 % (5-44) | 21 % (13-34) | 26 % (20-36) | 49 % (40-60) | 75 % (65-87) |
| AMAZONAS | 12 % (3-53) | 7 % (2-22) | 8 % (5-16) | 21 % (13-35) | 31 % (22-43) | 64 % (47-90) |
| ANTIOQUIA | 22 % (6-79) | 29 % (10-85) | 35 % (21-57) | 38 % (28-51) | 60 % (48-73) | 81 % (70-94) |
| ARAUCA | 12 % (3-49) | 9 % (3-28) | 12 % (7-23) | 22 % (14-35) | 36 % (26-50) | 87 % (63-100) |
| ATLANTICO | 7 % (2-27) | 7 % (2-22) | 12 % (7-19) | 18 % (13-24) | 35 % (28-44) | 62 % (53-72) |
| BARRANQUILLA | 8 % (2-30) | 10 % (3-30) | 15 % (9-25) | 22 % (16-30) | 46 % (37-57) | 67 % (57-78) |
| BOGOTA | 18 % (5-65) | 19 % (6-56) | 25 % (15-41) | 31 % (23-43) | 55 % (45-67) | 78 % (67-89) |
| BOLIVAR | 8 % (2-30) | 7 % (2-21) | 19 % (10-35) | 23 % (16-33) | 52 % (39-70) | 72 % (59-89) |
| BOYACA | 15 % (4-58) | 18 % (6-55) | 29 % (17-50) | 31 % (22-43) | 64 % (50-82) | 84 % (70-100) |
| CALDAS | 20 % (5-76) | 46 % (14-100) | 32 % (19-56) | 41 % (29-57) | 73 % (57-93) | 100 % (84-100) |
| CAQUETA | 8 % (2-30) | 14 % (4-43) | 12 % (7-21) | 17 % (12-23) | 40 % (32-52) | 74 % (61-90) |
| CARTAGENA | 17 % (4-62) | 17 % (6-51) | 29 % (17-49) | 40 % (29-56) | 73 % (57-93) | 89 % (74-100) |
| CASANARE | 12 % (3-49) | 13 % (4-41) | 13 % (7-24) | 25 % (17-39) | 39 % (28-53) | 79 % (59-100) |
| CAUCA | 13 % (4-51) | 16 % (5-51) | 27 % (15-48) | 28 % (20-40) | 50 % (39-63) | 84 % (70-100) |
| CESAR | 7 % (2-27) | 12 % (4-35) | 16 % (10-28) | 24 % (17-33) | 44 % (35-54) | 75 % (63-89) |
| CHOCO | 5 % (1-21) | 6 % (2-20) | 11 % (6-21) | 17 % (11-25) | 53 % (37-78) | 78 % (59-100) |
| CORDOBA | 5 % (1-17) | 6 % (2-18) | 11 % (7-18) | 18 % (13-24) | 37 % (30-46) | 61 % (52-71) |
| CUNDINAMARCA | 15 % (4-55) | 17 % (6-50) | 21 % (13-35) | 23 % (17-32) | 47 % (37-58) | 79 % (67-93) |
| GUAINIA | 10 % (2-49) | 14 % (3-71) | 12 % (5-29) | 28 % (12-68) | 100 % (41-100) | 92 % (43-100) |
| GUAJIRA | 7 % (2-27) | 7 % (2-21) | 13 % (7-22) | 21 % (15-30) | 44 % (34-56) | 68 % (55-82) |
| GUAVIARE | 46 % (6-100) | 10 % (3-38) | 100 % (19-100) | 17 % (10-32) | 40 % (24-71) | 80 % (50-100) |
| HUILA | 9 % (2-34) | 13 % (4-38) | 16 % (10-27) | 22 % (16-31) | 46 % (36-57) | 69 % (58-81) |
| MAGDALENA | 4 % (1-14) | 6 % (2-17) | 9 % (5-15) | 13 % (10-19) | 33 % (26-43) | 50 % (42-60) |
| META | 17 % (4-63) | 17 % (6-52) | 18 % (11-31) | 23 % (16-32) | 47 % (37-60) | 79 % (66-94) |
| NARIÑO | 9 % (2-34) | 11 % (4-33) | 19 % (11-33) | 24 % (17-33) | 43 % (34-54) | 70 % (59-82) |
| NORTE SANTANDER | 6 % (2-23) | 6 % (2-18) | 9 % (5-15) | 14 % (10-19) | 31 % (25-38) | 60 % (51-70) |
| PUTUMAYO | 9 % (2-34) | 7 % (2-21) | 10 % (6-18) | 14 % (10-21) | 29 % (22-39) | 62 % (49-79) |
| QUINDIO | 20 % (5-78) | 20 % (7-63) | 29 % (17-51) | 36 % (25-50) | 48 % (38-61) | 99 % (82-100) |
| RISARALDA | 19 % (5-71) | 28 % (9-86) | 25 % (15-43) | 31 % (22-43) | 58 % (46-73) | 89 % (74-100) |
| SAN ANDRES | 23 % (4-100) | 100 % (100-100) | 69 % (22-100) | 29 % (17-48) | 100 % (85-100) | 100 % (93-100) |
| SANTANDER | 12 % (3-45) | 14 % (5-42) | 15 % (9-25) | 21 % (15-28) | 43 % (35-54) | 77 % (66-89) |
| STA MARTA D.E. | 10 % (3-38) | 8 % (3-24) | 16 % (9-27) | 23 % (16-33) | 46 % (36-59) | 81 % (66-100) |
| SUCRE | 11 % (3-41) | 11 % (4-33) | 17 % (10-29) | 22 % (16-31) | 50 % (40-64) | 77 % (65-92) |
| TOLIMA | 14 % (4-51) | 22 % (7-68) | 22 % (13-37) | 26 % (19-35) | 51 % (41-64) | 75 % (64-88) |
| VALLE | 16 % (4-57) | 15 % (5-44) | 21 % (13-35) | 25 % (18-33) | 44 % (36-54) | 68 % (59-79) |
| VAUPES | 12 % (2-67) | 100 % (100-100) | 81 % (14-100) | 29 % (14-62) | 100 % (59-100) | 100 % (58-100) |
| VICHADA | 100 % (100-100) | 100 % (100-100) | 100 % (100-100) | 21 % (10-44) | 50 % (24-100) | 100 % (54-100) |

As the pandemic deepened during the year, the number of tests as well as the number of positive Covid-19 increased. The evolution of the percentage of cases reported for Colombia and some its regions during the year 2020 is presented in fig. 5. Besides the pandemic outbreak date being different in each region, we consider data for a department or city only after it has reported its first 40 fatal cases, resulting in different initial dates in the time axis for different regions. Due to the delay between onset-to-death, in most of the regions already a number of cases had been registered before the first death was reported, explaining the initial decay during the first weeks of the pandemic of the percentage of cases reported. Afterwards, the most important feature in all cases is the steady rising during the year of the percentage of reporting, yet reaching beyond the 50% only in a few regions, and the narrowing of the confidence intervals as the number of cases reported increased. This behavior goes hand in hand with the rising in the number of daily test for Covid-19 in the country during that period. Colombian testing capability has been insufficient during the pandemic, with an accumulated positivity rate of 23% and a daily percent positive that, except during the first two months of the pandemic, has remained above 10% [18]. Nonetheless, the country has increased its testing capacity considerably, going from 2000 daily tests at the end of March to being able to perform around 45000 in July and more than 50000 by the end of 2020. It can be seen that by the last week of April the percentage of cases reported in Colombia starts to rise from 22% to reach 43% by the end of December 2020. The observed plateau between June and August coincides with a peak of positive cases reported in Colombia. Even though the number of daily tests also incremented on those dates, its rate did not match the rate at which new cases and, more importantly, new deaths rose. The effect of this peak of detected cases is apparent in almost all regions, where after September a continuous increment in the percentage of report is noticeable.

**FIG. 5.**
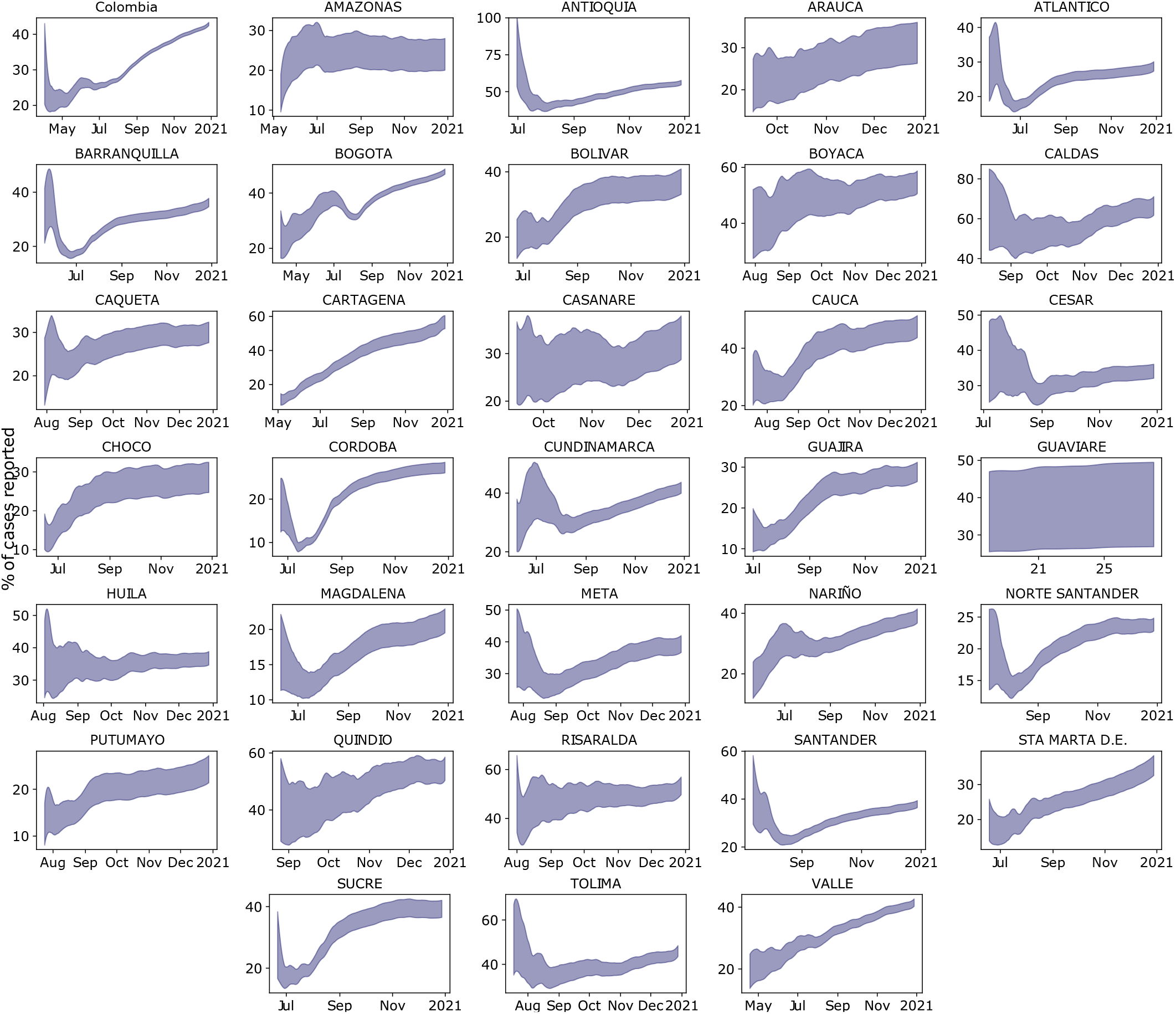
Evolution of the percentage of cases reported for Colombia and some its regions during the year 2020. For each region the 95% confidence interval of age-aggregated percentage of reported cases is shown.

At the level of regions, our results show correlations with local public policies that had being carried out. Some examples can be clearly identify from fig. 5. For instance, early on in the pandemic Boyacá closed its borders [19], reopening them only during the second semester of 2020. This caused a surge in the number of cases by the end of October and beginning of November. Nonetheless, despite the pronounced peak in the number of cases, the testing capacity was enough to keep level the reporting percentage over the course of those months. Amazonas, the largest department in Colombia by area and one of the less populated, constitutes another interesting example. It hit the headlines several times because it had the largest number of deaths per inhabitant at the beginning of the pandemic [20]. As a consequence, the number of test in the region was increased to the point that it became the region with more performed tests per inhabitant in the country. The percentage of cases report sharply increased during these early months, only to stagnate in July at a level well below the country’s average.

## IV. DISCUSSION

Our results present a detailed picture of how the Covid-19 infection has been tracked in Colombia during the year 2020. Yet, before commenting on some of the issues that they rise, a few words on their stability are called for. We notice that the figures in our analysis are sensitive to the chosen baseline CFR. For this work we selected the data reported by the Republic of Korea [16] (see also Table II), as this country tracking of the Covid-19 infections for the time span studied was considered thorough. A selection of a different benchmark country should lead to changes in the values present here, nevertheless, we expect the observed trends to remain the same. Furthermore, taking into account Colombia’s demographics should damps those effects. This, as we noticed before, yields, for example, a lower blCFR for Colombia as compared to the one of the Republic of Korea.

The inclusion of the country’s demographics to estimate its CFR and those of its regions is the most relevant feature of our work. This allows us to deal with Covid-19’s mortality variability in different age-groups, arguably one of the main factors driving the pandemic, and to account for its effects in the sub-report estimation. In particular, the larger vulnerability of older population compared to younger population explains the observed tendency of Colombia’s departments with a younger populations towards a lower blCFR than the country’s value (see Table III). Its effects on the corrected CFR and the percentage of cases reported, though less intuitive, are appreciable too. Despite the testing strategies implemented in Colombia not having uniformly sampled the population [18], the majority of it has been done on the population under 50 years old. This notwithstanding, and contrary to the expectations, there is a higher percentage of cases reported for the population with 70 years or more, and a noticeable gap between it and the percentage of report in the younger population. The higher vulnerability of the older age-groups helps explaining this behaviour too. Firstly, as we already clarified, for these age groups the blCFR is larger. Secondly, contrary to what happens to the younger population, in which most of the infected individuals only develop mild symptoms [21, 22] and many of the cases pass undetected, in the older population most of the infected individuals present clear symptoms, are therefore tested and their cases detected.

To conclude, we remark that our results are qualitatively consistent with the ones reported by Russell *et al*. [12] for Colombia. The differences in scales both in time and percentage of report are due to the different choice on benchmarks. While our delay density distribution *f* (*t*) is modeled from the the onset-to-death data for Colombia, in their work they use hospitalisation-to-death data from Wuhan [23], which may explain the shift in time of the first peak of reporting. They also use the CFR of Covid-19 for China [23], whereas our accounting of the country’s demographics, noticing that Colombia’s population is younger, leads to a lower baseline CFR, ultimately leading to our lower estimation of the percentage of cases reported.

## V. LIMITATIONS

Our study relays on some assumptions that limit the reach of our results:

- All of our estimations are linked and biased by the CFR of the benchmark country. In particular, our determination of the blCFR is very sensitive to it. Hence, small changes in the benchmark CFR can lead to considerable changes in the percentage of cases reported.
- We assume under-reporting is the only ground for the deviation of the CFR from the baseline CFR. A more comprehensive approach should consider additional factors, like the burden of the health-care system and other socio-economical factors that can influence the capacity of a country or region to track the Covid-19 pandemic.
- We do not account for comorbidities of the population which can influence the vulnerability (i.e. the baseline CFR) of a country or a region [24].
- Our results ignore fatal cases under-counting. A factor that could be relevant for the case of Colombia [25].
- We do not take into account different strains of Covid-19 and the different CFRs for each type [26, 27].

## VI. CONCLUSIONS

In this study we present a comprehensive analysis of the percentage of Covid-19 cases reported in Colombia, providing a clearer picture of the magnitude of the pandemic in this country as of December 28, 2020. Specifically, we estimate that the 1’594.497 of confirmed positive cases reported during 2020 represents only the 44% of the total number of infected people in Colombia during this period. In Bogotá, which accounts for around a third of the reported cases in the country, the percentage of report was only of 48%.

Our estimation accounts for both the time delay between symptom’s onset and death caused by the Covid-19 disease, and the country’s, its regions’ and cities’ demographics in order to estimate their local CFR. It is worth stressing, that this last feature in our method provides it with a sensitive account of the age-dependent effects of Covid-19, not present, for instance, in [12], which allows us to study the under-reporting of cases in each of the regions of the country accounting for their demographic particularities. Thus, we were able to identify more vulnerable departments, having a large CFR, from those expected to have a lower mortality.

We show that the overall value of the percentage of cases reported by the country conceals the large inhomogeneity of its efforts to trace the infection in its territory, with a marked tendency for departments lying outside the Andean region to have a lower percentage of report. We provide a twofold explanation for the observed sub-report differences in Colombia’s regions. The first relevant factor was the country’s policy to track the infection. In it, much of the Covid-19 testing was concentrated in the Andean region, where the largest fraction of the population lives, leaving the departments in the periphery with smaller testing programs. The second factor is less evident and closely linked with the regions demographics. Due to the greater vulnerability of the older population against Covid-19, the CFR for this group is larger than for other groups in the population, making the percentage of reported cases for older population in all regions to be significantly higher than that for the younger population.Therefore, in departments far from the center of the country, where older age-groups correspond to smaller fractions of the population, this factor pushes the percentage of report towards lower values.

Finally, our results could be of use to contrast with seroprevalence studies for Colombia during the 2020 [28, 29] in order to estimate the real size of the pandemic in the country. A relevant information in the present stage for the planning of the vaccination program of the Country.

## Data Availability

We developed a Python library that eases the extraction, load and transformation of raw data from the INS database. The code is freely available at https://gitlab.com/hubrain/covid19. Also, we designed a dashboard were we keep a daily record of how reporting percentages are changing at each region which can be found at http://covid19.hubrain.co. Furthermore, the code to reproduce the figures and tables here presented is in https://gitlab.com/hubrain/covid19-paper.

## ACKNOWLEDGMENTS

We kindly acknowledge stimulating discussions with Luis Fernando García Moreno, Marcela Henao Tamayo, and Rafael Hurtado.

## DATA AVAILABILITY

We developed a Python library that eases the extraction, load and transformation of raw data from the INS database. The code is freely available at https://gitlab.com/hubrain/covid19. We also maintain a dashboard, where a daily record of how percentage of cases reported is changing in Colombia and its regions, at http://covid19.hubrain.co. The code to reproduce the figures and tables presented in this document can be found at https://gitlab.com/hubrain/covid19-paper.

